# Multicenter Evaluation of BACT-Info. and an Infection Algorithm Using Urine Flow Cytometry among Clinically Diagnosed UTI Patients in Indonesia

**DOI:** 10.64898/2025.12.07.25341799

**Authors:** Andaru Dahesihdewi, Tonny Loho, Aryati, Ria Triwardhani, Muji Rahayu, Ika Priatni, Dewi Lesthiowati, Ni Sayu Dewi, Yosanti Elsa Kartikawati, M. I. Diah Pramudianti, Brigitte Rina Aninda Sidharta, Leli Saptawati, Ferdy Royland Marpaung, Hartono Kahar, Asvin Nurulita, Rachmawati A. Muhiddin, Anti Dharmayanti, Luci Liana, Sianny Herawati, I Nyoman Wande, Riat El Khair, Ira Puspitawati, Hani Susianti, Agustin Iskandar

## Abstract

Urinary tract infections (UTIs) are the most common infections in both outpatient and inpatient settings, contributing significantly to morbidity, reduced quality of life, and antimicrobial overuse. Although urine culture remains the diagnostic gold standard, it poses practical limitations in clinical workflows. Rapid diagnostic methods such as urine flow cytometry (UF) offer potential for timely, reliable UTI detection.

We conducted a multicenter diagnostic study to evaluate the performance of BACT-Info. and UTI-Info. flags on the UF-5000/4000 system in detecting UTIs, using presumptive Gram staining and urine culture (>10⁵ CFU/mL) as references.

A total of 763 patients with suspected UTI were enrolled, and 721 patients—with uropathogenic bacteria and complete data—were included in the final analysis (384 with culture-confirmed UTI and 337 without UTI). The diagnostic value of nitrituria was highly specific, while leukocyte esterase was sensitive. For BACT-count and WBC-count of UF-5000/4000, the AUCs were 0.85 and 0.69, respectively. Using cutoffs of WBC >82.05/µL or bacteria >975.4/µL, the UTI-Info flag demonstrated 89% sensitivity, 54% specificity, 69% positive predictive value, and 82% negative predictive value. Positive and negative likelihood ratios were 1.93 and 0.20, respectively. The agreement between the BACT-Info. flag and Gram typing showed Kappa values of 0.716 for Gram-negative and 0.216 for Gram-positive bacteria when compared to culture, and 0.721 and 0.401, respectively, when compared to presumptive Gram staining.

The UF-5000/4000 UTI-Info. and BACT-Info. flags show promise as a rapid UTI screening tool. The combination of these flags with urinalysis parameters, nitrite testing and leukocyte has potential to be developed into a diagnostic algorithm for early and sensitive UTI prediction. Such an approach may reduce unnecessary urine cultures and support timely, appropriate empiric antibiotic therapy. Establishing optimal cutoffs tailored to specific clinical settings is essential to enhance diagnostic accuracy and improve clinical utility.

## Introduction

Urinary tract infection (UTI) is a prevalent infection in developing countries such as Indonesia, where the prevalence ranges from 5% to 15%, with an annual incidence of 90–100 cases per 100,000 population [1]. UTIs primarily result from bacterial invasion of the urinary tract, commonly originating from the perineal or perianal area and entering the urethra in a retrograde manner. The infection can involve various parts of the urinary system, but the urinary bladder (cystitis) is commonly affected.

Women are disproportionately affected due to anatomical and physiological factors, including a shorter urethra, proximity of the urethral orifice to the anus, hormonal changes during menopause, and the use of certain contraceptives such as vaginal diaphragms and spermicides. Other risk factors include urinary catheterization, structural abnormalities of the urinary tract, diabetes mellitus, and immunosuppression [2].

Antibiotics are the mainstay of UTI treatment. However, inappropriate or excessive use of antibiotics contributes significantly to the growing problem of antimicrobial resistance (AMR), which has emerged as a major global health threat. AMR is associated with increased treatment failure, complications, longer hospital stays, and higher healthcare costs [3]. Overprescribing antibiotics, particularly in cases without confirmed bacterial infection, not only contributes to resistance but also exposes patients to unnecessary side effects and increases the likelihood of recurrent infections [4].

Accurate diagnosis is essential for appropriate antibiotic use. Urine culture is considered the gold standard for diagnosing UTI and is a common microbiological culture requested in laboratories [5]. However, it is time-consuming, labor-intensive, and often unavailable in resource-limited settings. Furthermore, studies show that up to 60% of urine cultures from suspected UTI cases yield negative results, indicating a substantial burden on healthcare resources with limited diagnostic yield [6].

Given these challenges, there is a critical need for a rapid, reliable, and cost-effective method to identify patients who truly require urine culture testing. Such an approach would support antimicrobial stewardship, enhance laboratory efficiency, reduce unnecessary antibiotic use, and ultimately help curb the rise of AMR.

In this study, we evaluated the performance of an automated urinalysis system using flow cytometry to screen patients with clinically suspected UTI for the need of urine culture. Conducted as a multicenter study across several hospitals in Indonesia, the investigation focused on the system’s ability to provide rapid, quantitative bacteriuria data alongside Gram classification through two integrated features: UTI-Info. and BACT-Info.. These flags in Sysmex urinalysis analyzer help identify the likelihood of a urinary tract infection and indicate the type of bacteria (positive, negative, combination or unspecified Gram-type differentiation), respectively. They support more targeted and efficient diagnostic decision-making. The results of this study are also aimed at developing an algorithm that integrates urinalysis and urine flow cytometry using the Sysmex UF-5000/4000 for UTI screening, in order to optimize the need for urine culture and guide empirical antibiotic therapy.

## Materials and Methods

### Study Design

This was a multicenter, cross-sectional diagnostic study. The performance of the BACT-Info. or UTI-Info. flag from automated urinalysis was evaluated against presumptive Gram staining and urine culture, performed independently and in a blinded manner.

Based on a study by Ren et al., which evaluated the Fully Automated Urine Particle Analyzer UF-5000 to rapidly distinguish culture-negative urine specimens in patients with suspected UTI in an Asian population, the sensitivity and specificity of the UTI-Info. flag were reported as 97.8% and 74.6%, respectively [7]. Assuming a UTI prevalence of 5% in the Indonesian population [1], the minimum required sample size was 662 to meet both sensitivity and specificity objectives. To account for potential data loss, the target was increased to 700.

Eligible participants were adults (≥18 years) with clinically suspected UTIs, either hospitalized or attending outpatient clinics. Urine specimens were collected via clean-catch midstream or aseptic catheterization using sterile containers. Exclusion criteria included menstruation, pregnancy, and samples with poor collection quality (e.g. delayed transport, contamination, or insufficient volume).

### Study Location and Duration

Human participants were prospectively recruited for this study. The recruitment period began on 4 March 2023 and ended on 4 September 2023. The study was conducted across 10 hospitals in Indonesia, recruiting 763 patients with suspected UTIs (Fig 1).

**Figure 1.**
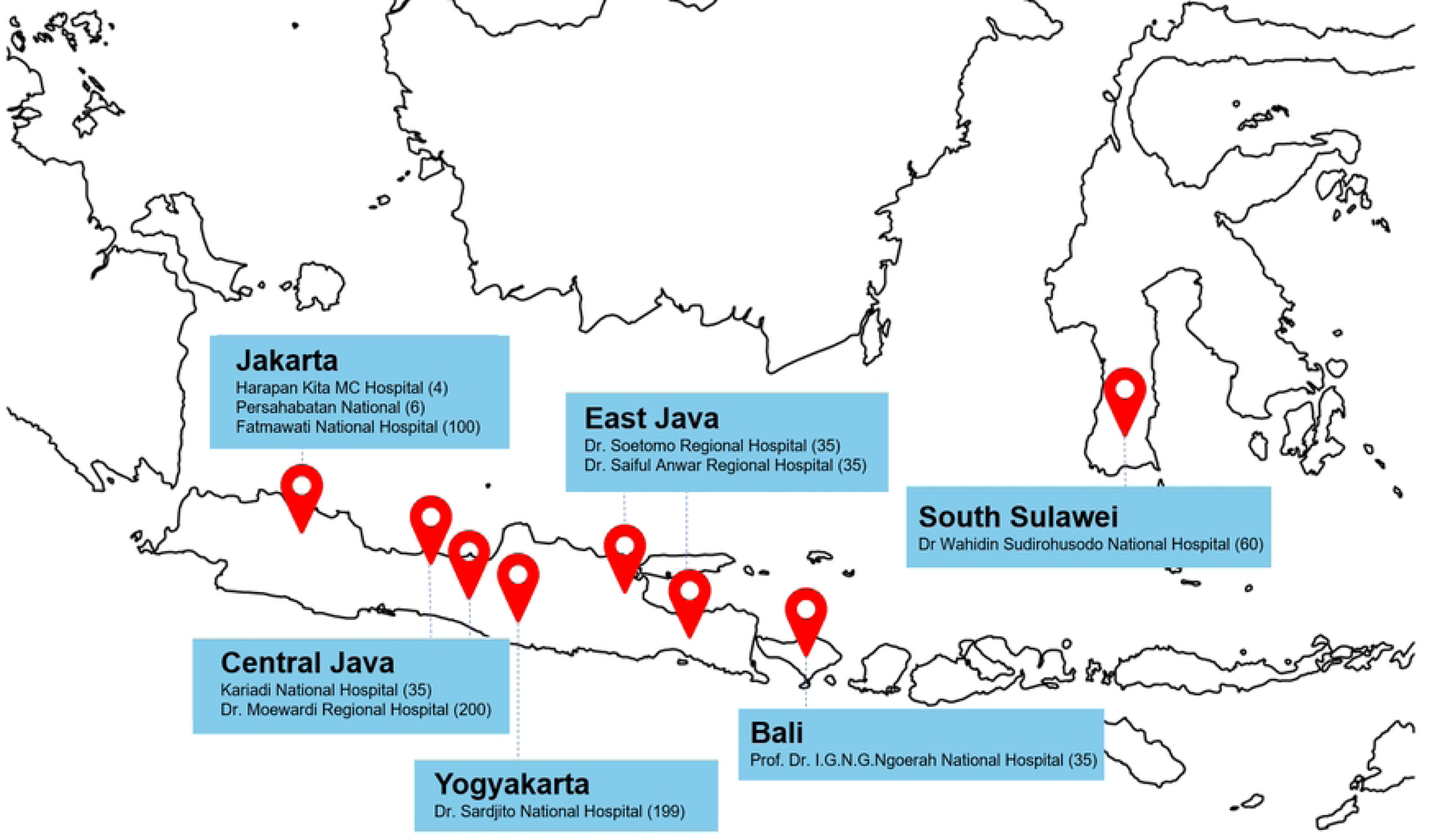
Geographic Distribution of Participating Hospital Sites in Indonesia. Map showing the distribution of patients enrolled from various participating hospitals across Indonesia, including facilities in Jakarta, Central Java, Yogyakarta, East Java, Bali, and South Sulawesi. The numbers in parentheses represent the number of patients recruited from each hospital

### Data Collection and Laboratory Methods

Urine samples were analyzed using the Fully Automated Urine Particle Analyzer Sysmex UF-5000/4000 within two hours of collection. The index tests included BACT-Info. results (Gram Positive?, Gram Negative?, Gram Pos/Neg?, Unclassified, or no flag) and UTI-Info. results (UTI or non-UTI). Reference standards were the presumptive Gram stain (Gram-positive, Gram-negative, mixed, or no bacteria found) and urine culture, reported as positive or negative for UTI. UTI was defined by urine culture resulted (≥10⁵ CFU(Colony Forming Unit)/mL for midstream/catheter samples; >10² CFU/mL for straight catheter samples), along with Gram type, and bacterial species. Laboratory analysis included routine urinalysis, urine flow cytometry, and urine culture. Urinalysis provided semiquantitative measurements such as white blood cells and nitrite. Flow cytometry generated semiquantitative and categorical data on white blood cells, red blood cells, epithelial cells, casts, sperm, yeast, mucus, and bacterial counts. Urine culture supplied quantitative CFU results along with bacterial Gram type and species identification.

Additional variables collected included age, gender, UTI symptoms (e.g., dysuria, fever, hematuria), history and classification of UTI (recurrent/new, complicated/uncomplicated, upper/lower UTI); catheterization status categorized as none, short term (0-14 days), long term (more than 14 days), or recurrent; urinary tract abnormalities; immune status, including menopause, immunosuppressive therapy.

### Data Analysis

Categorical variables were presented as frequencies and percentages. Normally distributed numerical data were expressed as mean ± standard deviation (SD), while non-normally distributed data were reported as median with minimum and maximum values. Statistical comparisons for categorical data were conducted using the Chi-square test or Fisher’s exact test, while numerical data were analyzed using the independent t-test for normally distributed variables, the Mann-Whitney U test for non-normally distributed data, and ANOVA when comparing more than two groups. Diagnostic accuracy was assessed using receiver operating characteristic (ROC) curves and 2×2 tables to compare UTI-Info. results with culture findings. Diagnostic assessment presented as sensitivity, specificity, positive predictive value (PPV), negative predictive value (NPV), likelihood ratios, and overall accuracy. while the agreement between BACT-Info. and Gram stain or culture results were estimated using the Kappa statistic. Data analysis was performed using Microsoft Excel (Office 365), MedCalc, and SPSS version 20.

### Ethics Statement

All participants provided written informed consent prior to enrollment. This study received ethical approval from the Ethics Committee of the Faculty of Medicine, Public Health, and Nursing, Universitas Gadjah Mada/Dr. Sardjito Hospital, Yogyakarta (Ref: KE/FK/1203/EC/2022). All patient data were anonymized and handled with strict confidentiality.

## Results

### Study population and characteristics of the participants

This study analyzed urine samples from 763 patients with suspected UTIs across 10 major referral hospitals in Indonesia (Fig 2). The largest shares came from Dr. Moewardi Surakarta (200 patients; 26.2%), Dr. Sardjito Yogyakarta (199; 26.1%), and Fatmawati Jakarta hospitals (100; 13.1%), while the remaining seven hospitals contributed 4–60 patients each (34.6%). Most patients were female (Table 1). The sample size was adequate for assessing UF diagnostic validity, but the concentration of samples (66%) from three hospitals in Jakarta, Central Java, and DI Yogyakarta may introduce selection bias, limiting generalizability.

**Figure 2.**
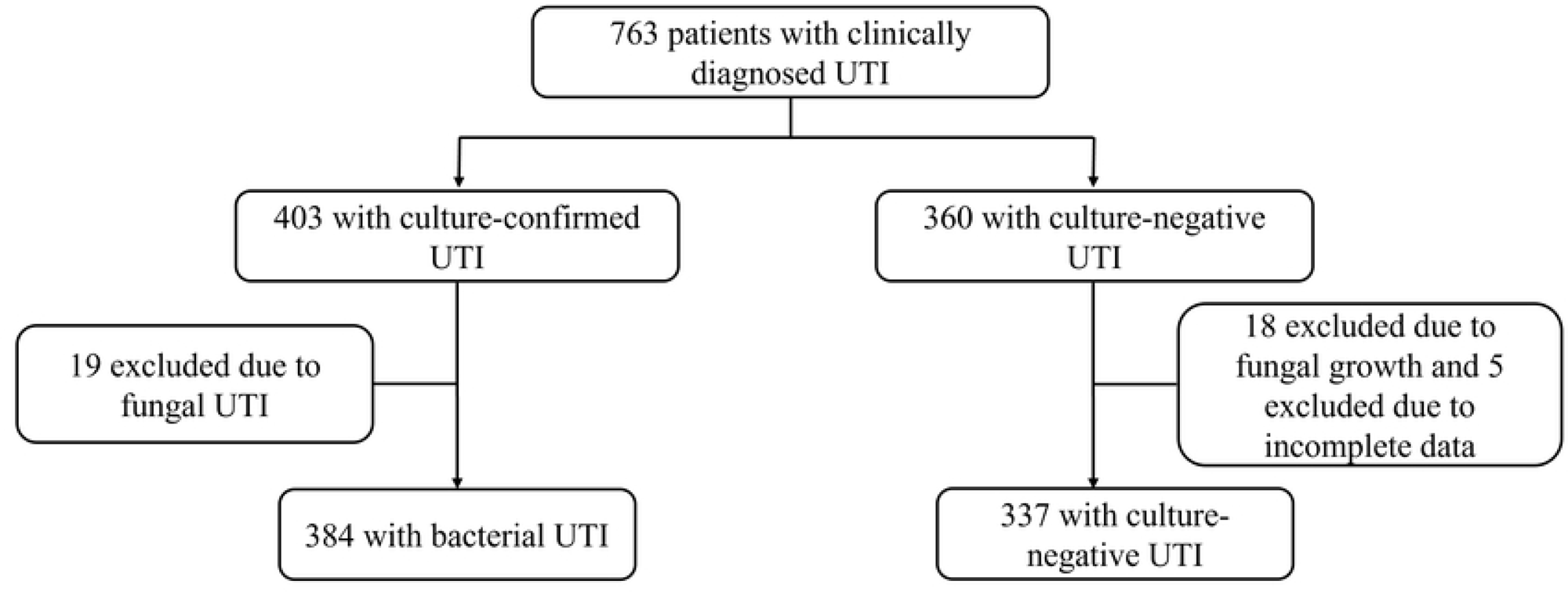
Patient enrollment and selection flow for UTI Diagnostic Validity Analysis. A total of 763 clinically suspected UTI patients were initially enrolled. Among them, 403 cases were confirmed as UTI based on urine culture, while 360 were culture-negative. Forty-two patients were excluded due to fungal culture results or incomplete data, resulting in 721 eligible subjects for subsequent diagnostic validity analysis—comprising 384 culture-confirmed bacterial UTI cases and 337 non-UTI cases.

**Table 1.** Patient distributions, their age and sex based on their admitting hospital.

| Hospital | n (%) | Age (y.o.) |  | Sex |  |  |
| --- | --- | --- | --- | --- | --- | --- |
|  |  | Mean ± SD | *p* * | Male<br>n (%) | Female<br>n (%) | *p* ** |
| Harapan Kita Hospital, Jakarta | 4 (0.52) | 36.0 ± 5.7 | <0.001 | 0 | 4 (0.9%) | <0.001 |
| Persahabatan Hospital, Jakarta | 60 (7.86) | 53.5 ± 16.5 |  | 21 (7.1) | 39 (8.4%) |  |
| Fatmawati Hospital, Jakarta | 100 (13.10) | 47 ± 12.4 |  | 40 (13.5%) | 60 (12.8%) |  |
| Dr Kariadi Hospital, Semarang | 35 (4.59) | 48.5 ± 17.5 |  | 5 (1.7%) | 30 (6.4%) |  |
| Dr. Moewardi Hospital, Surakarta | 200 (26.22) | 46.2 ± 23.1 |  | 74 (25%) | 126 (27%) |  |
| Dr. Sardjito Hospital, Yogyakarta | 199 (26.08) | 53.3 ± 17.1 |  | 88 (29.7%) | 111 (23.8%) |  |
| Dr. Saiful Anwar Hospital, Malang | 35 (4.59) | 54.2 ± 16.4 |  | 11 (3.7%) | 24 (5.1%) |  |
| Dr. Soetomo Hospital, Surabaya | 35 (4.59) | 48.8 ± 18 |  | 9 (3%) | 26 (5.6%) |  |
| Prof. Dr. I.G.N.G. Ngoerah Hospital, Denpasar | 35 (4.59) | 49 ± 16.9 |  | 7 (2.4%) | 28 (6%) |  |
| Dr. Wahidin Sudirohusodo Hospital, Makassar | 60 (7.86) | 50.4 ± 16.8 |  | 41 (13.9%) | 19 (4.1%) |  |
| **Total** | **763 (100)** |  |  | **296 (38.8%)** | **467 (61.2%)** |  |

\*ANOVA test; \*\*Fisher's exact test; *p*<0.05 indicates statistical significance; SD: standard deviation.

### Comparison of Patient Characteristics Based on UTI Status

Of the 763 patients, 403 (52.8%) met UTI criteria (Table 2). Participants were mostly female. UTI patients were significantly older (mean age 52.2 vs. 46.9 years, *p*<0.0001). Clinical symptoms significantly associated with UTI included dysuria, pollakiuria, nocturia, and hematuria (*p*<0.001). UTI patients also had higher rates of short-term catheter use and menopause (*p*=0.030 and *p*=0.002, respectively). Interestingly, immunosuppressant therapy was more common in non-UTI patients (*p*<0.001), while BMI was lower in UTI patients (*p*=0.020). Other variables, such as comorbidity, suprapubic pain, and fever, did not show significant differences between groups. Among UTI cases, 31% were non-complicated, 21.6% were complicated, and 33.7% were lower UTI. Recurrent UTIs made up 22.8% of cases. Only 10.4% UTI were classified as healthcare-associated infections, meaning the UTI occurred in the hospital after more than 48 hours of admission. A limitation of the study is that some patients’ clinical data could not be retrieved from medical records.

**Table 2.**
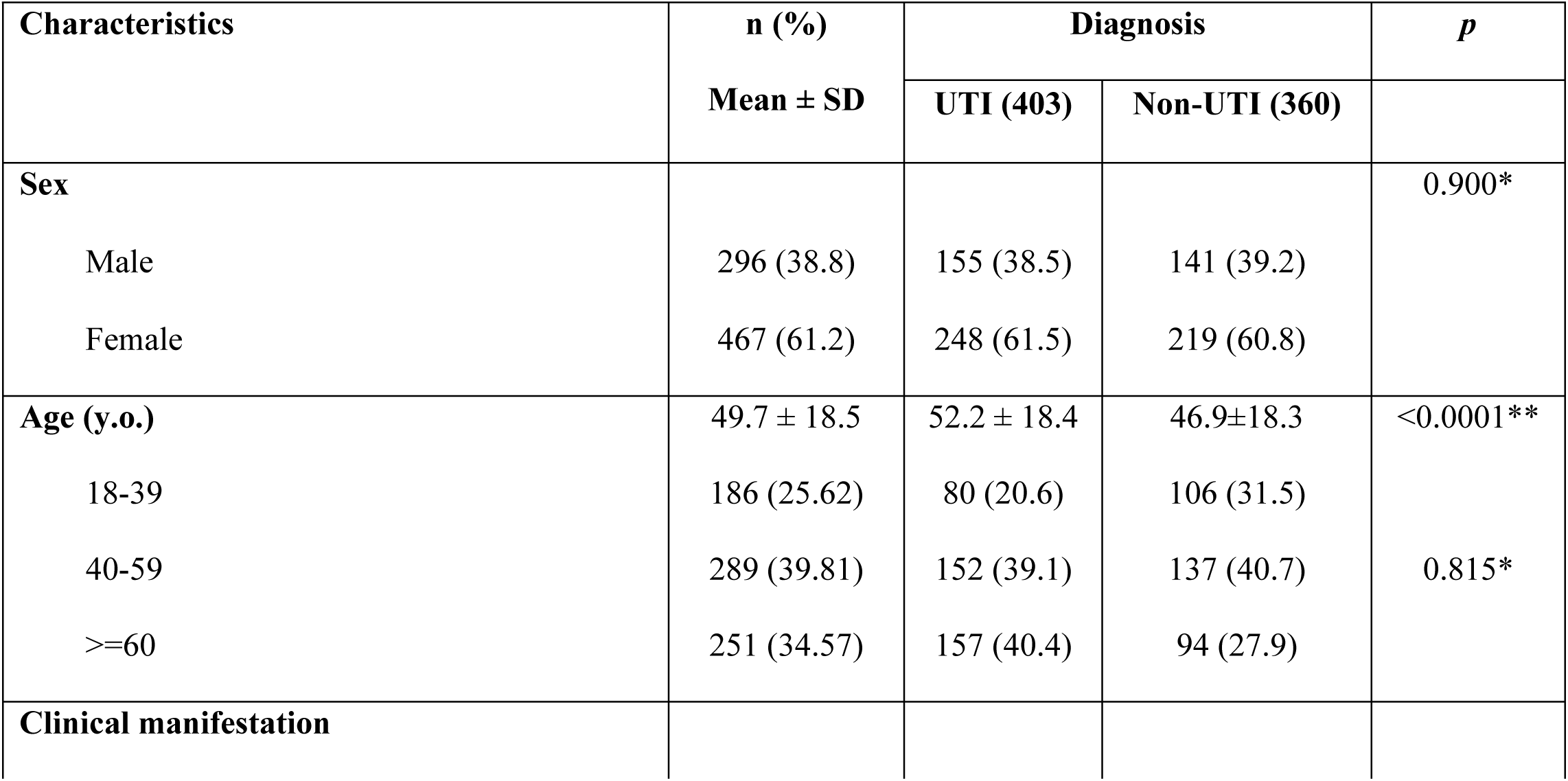
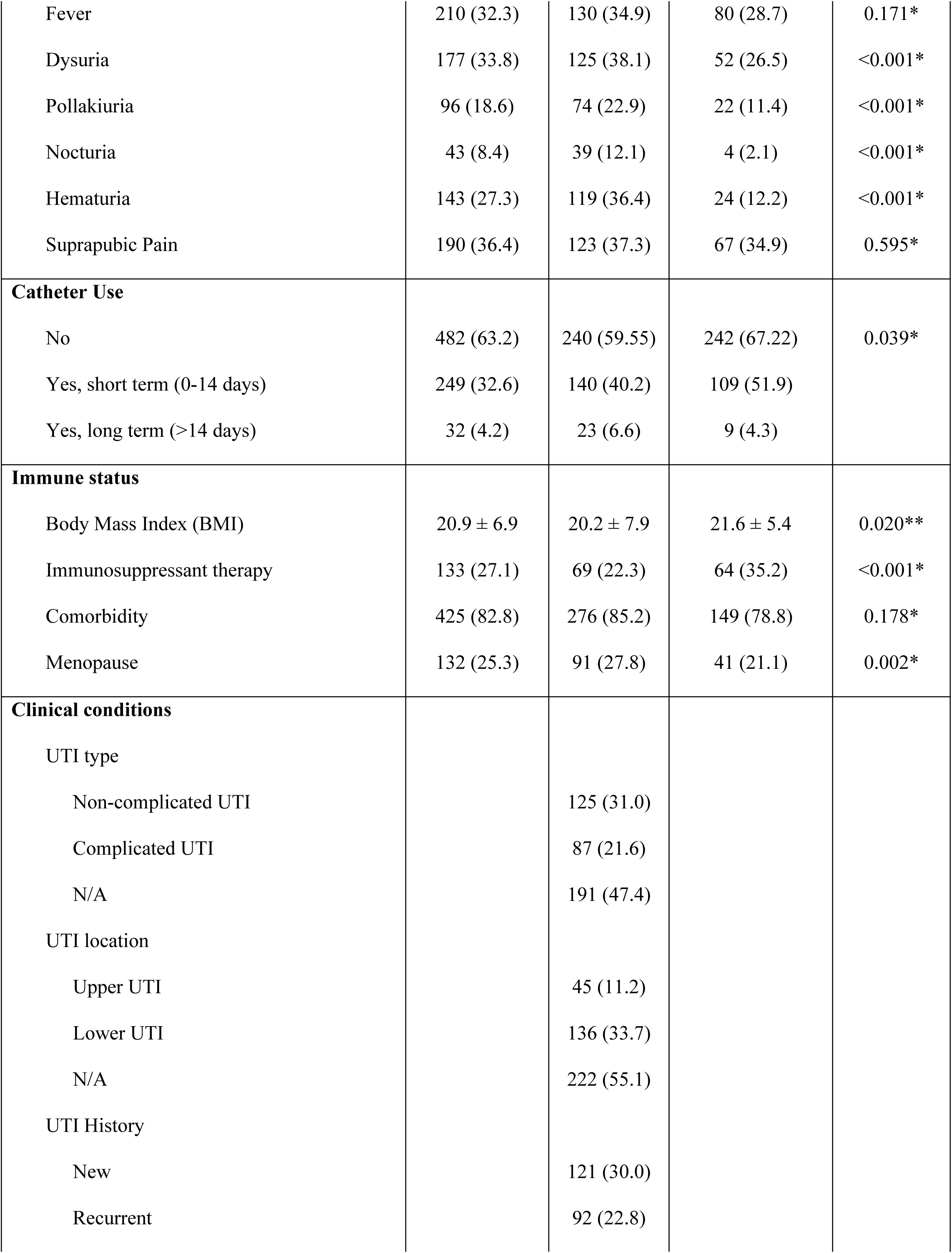
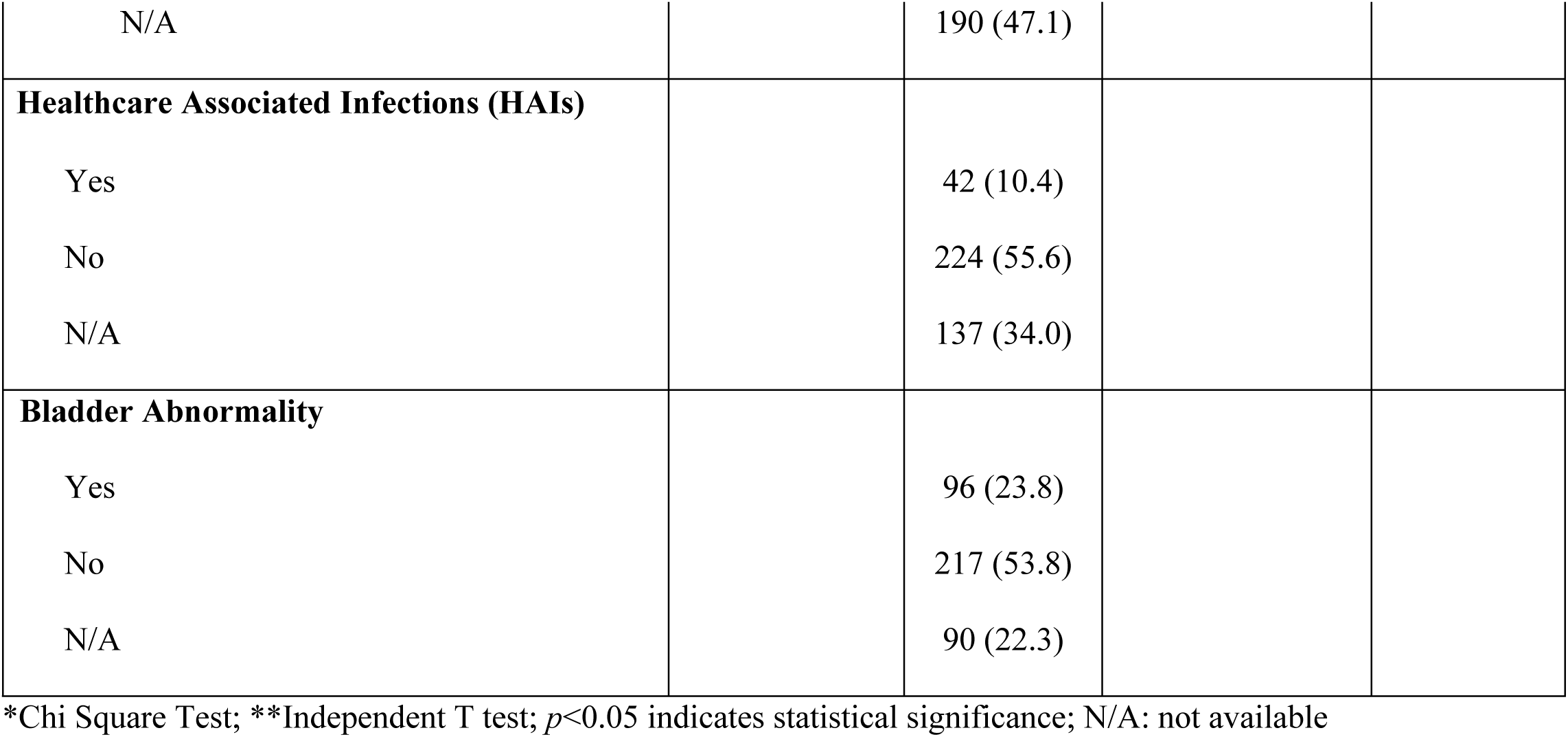
Patient characteristics based on UTI diagnosis (n=763)

### Urinalysis Profiles of Study Patients

The urinalysis results in this study’s subjects showed a significant difference between the UTI group and the non-UTI group in the parameters of nitrite, microscopic leukocytes, urinary casts, and bacteria. The results of other urinalysis parameters were relatively similar between UTI and non-UTI subjects. Overall, Table 3 illustrates the relationship between clinical data, the results of urine dipstick and microscopic sediment examination with UTI culture outcomes. The observed associations may support early indication considerations for infection based on urinalysis findings. For patients with positive urine culture, the identified microorganisms are listed in S1 Table 1.

**Table 3.** Laboratory Results with UTI-Info. Flag from UF-5000/4000.

| Urinalysis Laboratory Results | Total Patients<br>(n=763) | UTI-Info. Flag | | $p^{\#}$ |
| --- | --- | --- | --- | --- |
|  |  | Yes<br>(n=421) | No<br>(n=342) |  |

|  | n (%) | n (%) | n (%) |  |
| --- | --- | --- | --- | --- |
| **Color** |  |  |  |  |
| Clear/Transparent | 2 (0.3) | 1 (0.2) | 1 (0.3) | 0.267 |
| Brown | 39 (5.1) | 31 (7.4) | 8 (4.7) |  |
| Yellow | 238 (31.2) | 231 (54.9) | 7 (75.1) |  |
| Reddish/Pink | 357 (46.8) | 100 (23.8) | 257 (11.1) |  |
| Orange | 75 (9.8) | 37 (8.8) | 38 (6.7) |  |
| White and cloudy | 24 (3.1) | 1 (0.2) | 23 (0.0) |  |
| Others | 28 (3.7) | 20 (4.8) | 8 (2.0) |  |
| **Total** | **763 (100)** | **421 (100)** | **342 (100)** |  |
| **Blood** |  |  |  |  |
| (-) | 147 (19.3) | 55 (13.1) | 92 (26.9) | 0.021* |
| (-/+) | 37 (4.8) | 16 (3.8) | 21 (6.1) | 0.059 |
| 1+ | 84 (11.0) | 49 (11.6) | 35 (10.2) | 0.321 |
| 2+ | 125 (16.4) | 73 (17.3) | 52 (15.2) | 0.443 |
| 3+ | 370 (48.5) | 228 (54.2) | 142 (41.5) | 0.523 |
| **Total** | **763 (100)** | **421 (100)** | **342 (100)** |  |
| **Leukocyte esterase** |  |  |  |  |
| (-) | 217 (28.4) | 34 (8.1) | 183 (53.5) | 0.016* |
| 1+ | 86 (11.3) | 33 (7.8) | 53 (15.5) | 0.027* |
| 2+ | 156 (20.4) | 96 (22.8) | 60 (17.5) | 0.061 |
| 3+ | 304 (39.8) | 258 (61.3) | 46 (13.5) | 0.013* |
| **Total** | **763 (100)** | **421 (100)** | **342 (100)** |  |
| **Nitrite** |  |  |  |  |
| (-) | 613 (80.3) | 292 (69.4) | 321 (93.9) | 0.010* |
| (+) | 150 (19.7) | 129 (30.6) | 21 (6.1) |  |

| Total | 763 (100) | 421 (100) | 342 (100) |  |
| --- | --- | --- | --- | --- |
| Urinalysis Laboratory<br>Results and Urine<br>Sediment | Total Patients<br>n=763 | UTI-Info. Flag |  | *p*<sup>s</sup> |
|  |  | Yes<br>n=421 | No<br>n=342 |  |
|  | Median (min-max) | Median (min-max) | Median (min-max) |  |
| Density | 1.015 (1.001-1.090) | 1.014 (1.001-1.090) | 1.015 (1.002-1.044) | 0.428 |
| pH | 6.0 (5.0-9.0) | 6.0 (5.0-9.0) | 6.0 (5.0-9.0) | 1.000 |
| Flow Cytometry Urine<br>Sediment |  |  |  |  |
| Red Blood Cell<br>(RBC) | 56.8 (0-82381.2) | 76.15 (0.1-82381.2) | 34.2 (0-75958.8) | 0.613 |
| White Blood Cell<br>(WBC) | 224.1 (0-2534.937) | 438.3 (1.2-2534937) | 63.3 (0-438.689) | 0.023* |
| Urinary casts | 1.05 (0-352.23) | 1.54 (0-352.23) | 0.55 (0-181.56) | 0.032* |
| Bacteria | 1528.2 (0-2872.147) | 11.908 (0-2872.147) | 137.4 (0-1987.895) | 0.001* |

\**p*<0.05 indicates statistical significance. #Frequency data were analyzed using Chi-Square test. <sup>s</sup>Non-normally distributed data were analyzed using Mann-Whitney test.

### Association Between the UTI-Info. Flag and Bacterial Count in Urine Culture

The UF-5000/4000 flow cytometry showed a significant association between UTI-Info. flag results and urine colony counts. Among confirmed UTI cases, most samples had ≥100,000 CFU/mL, with 51.5% in that range and 11.1% exceeding 1 million CFU/mL. In contrast, 84.7% of non-UTI cases had bacterial counts below 100,000 CFU/mL. These findings indicate the system’s potential to differentiate between true UTI and non-UTI samples (*p*<0.05) (Table 4). In this analysis, 42 patients were excluded from the initial total patients of 763 due to fungal growth or incomplete data, resulting in 721 eligible subjects (Fig 2).

**Table 4.**
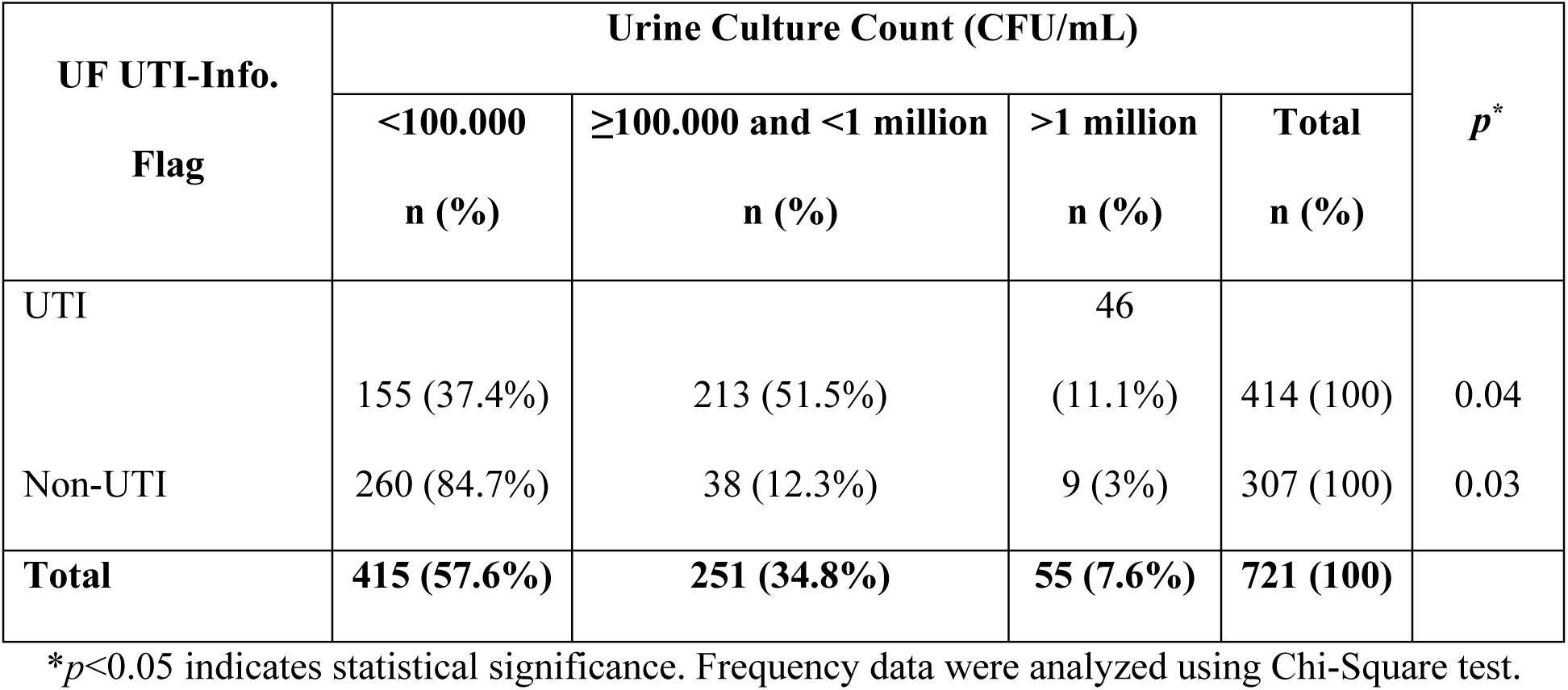
Detection Capability of UF-5000/4000 Flow Cytometry for UTIs Based on Urine Colony Count Results.

| UF UTI-Info.<br>Flag | Urine Culture Count (CFU/mL) | | | | $p^*$ |
| --- | --- | --- | --- | --- | --- |
| | <100.000<br>n (%) | $\geq 100.000$ and <1 million<br>n (%) | >1 million<br>n (%) | Total<br>n (%) | |
| UTI | 155 (37.4%) | 213 (51.5%) | 46 (11.1%) | 414 (100) | 0.04 |
| Non-UTI | 260 (84.7%) | 38 (12.3%) | 9 (3%) | 307 (100) | 0.03 |
| **Total** | **415 (57.6%)** | **251 (34.8%)** | **55 (7.6%)** | **721 (100)** |  |

\* $p<0.05$ indicates statistical significance. Frequency data were analyzed using Chi-Square test.

### Diagnostic Accuracy of UF-5000/4000 UTI-Info. Flag and Urinalysis Parameters in Differentiating UTI from Non-UTI Cases

The urinalysis results and UTI-Info. flag for UTI and non-UTI patients are shown in Tables 5-7. Urinalysis results showed that nitrite positivity was much higher in UTI patients (34.4%) compared to non-UTI (4.5%), indicating its usefulness as a good predictor. Leukocyte esterase levels were generally higher in UTI cases, with 53.9% showing 3+ results, versus 22.9% in non-UTI. Blood in urine was common in both groups, though slightly more concentrated in UTI patients. Mean white blood cell (WBC) and bacterial counts in urine sediment were markedly elevated in UTI patients (1273.1/µL and 29878.1/µL, respectively) compared to non-UTI (718.7/µL and 4550.5/µL), supporting their utility as diagnostic indicators (Table 5).

**Table 5.** Characteristics of Urine Dipstick and Sediment Examination Results among UTI and Non-UTI Culture Results (n=721)

| Test | Result | UTI (n=384)<br>n (%) | Non-UTI (n=337)<br>n (%) |
| --- | --- | --- | --- |
| **Nitrite (NIT)** | Positive (+) | 132 (34.4%) | 15 (4.5%) |
|  | Negative (-) | 252 (65.6%) | 322 (95.5%) |
| **Leukocyte Esterase (LEU)** | Negative (-) | 59 (15.4%) | 149 (38.8%) |
|  | 1+ | 32 (8.3%) | 41 (10.7%) |
|  | 2+ | 86 (22.4%) | 59 (15.4%) |
|  | 3+ | 207 (53.9%) | 88 (22.9%) |
| **Blood (BLD)** | Negative (-) | 65 (16.9%) | 78 (23.1%) |
|  | (+/-) | 19 (4.9%) | 16 (4.7%) |
|  | 1+ | 52 (13.5%) | 26 (7.7%) |
|  | 2+ | 61 (15.9%) | 55 (16.3%) |
|  | 3+ | 187 (48.7%) | 162 (48.1%) |
| **Sediment (Mean Values)** | WBC (/μL) | 1273.1± 362.1 | 718.7 ± 307.8 |
| | Bacteria (/μL) | 29878.1± 2904.9 | 4550.5 ± 1274.2 |

Urinalysis parameters showed varying diagnostic values for UTI detection among 721 patients (Table 6). Nitrite positivity had a high specificity of 95.5% but low sensitivity at 34.4%. Leukocyturia (>25/µL) and leukocyte esterase tests demonstrated high sensitivity (93.0% and 93.8%, respectively) but lower specificity (27.9% and 45.4%). Bacteriuria (>125/µL) also showed high sensitivity (90.9%) but limited specificity (39.2%). These results suggest that while some urinalysis markers are sensitive to detecting UTIs, their specificity varies.

**Table 6.**
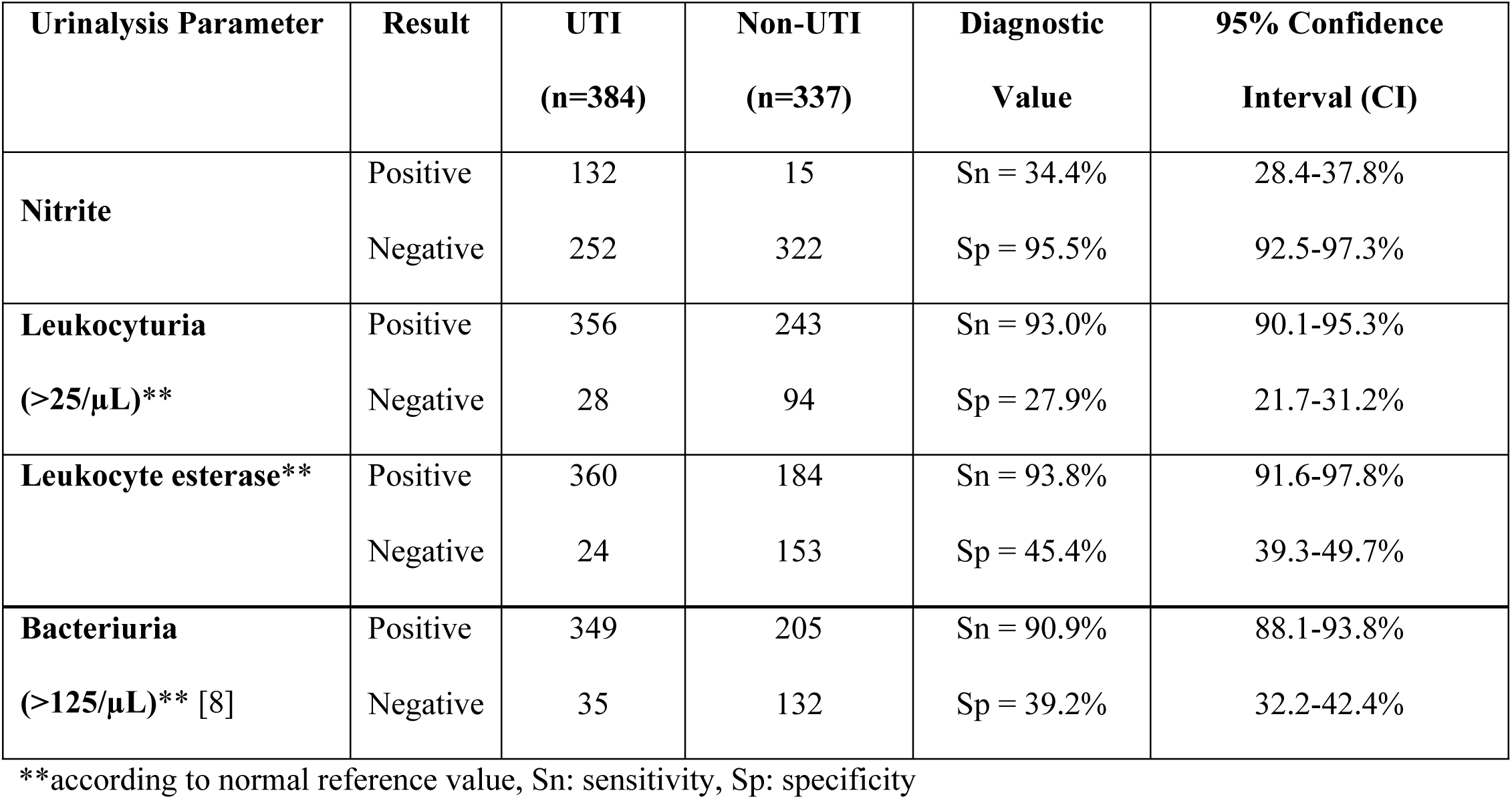
UTI Diagnostic Correlation with Urinalysis Results (n=721)

| Urinalysis Parameter | Result | UTI<br>(n=384) | Non-UTI<br>(n=337) | Diagnostic<br>Value | 95% Confidence<br>Interval (CI) |
| --- | --- | --- | --- | --- | --- |
| **Nitrite** | Positive | 132 | 15 | Sn = 34.4% | 28.4-37.8% |
|  | Negative | 252 | 322 | Sp = 95.5% | 92.5-97.3% |
| **Leukocyturia**<br>**(>25/μL)\*\*** | Positive | 356 | 243 | Sn = 93.0% | 90.1-95.3% |
|  | Negative | 28 | 94 | Sp = 27.9% | 21.7-31.2% |
| **Leukocyte esterase\*\*** | Positive | 360 | 184 | Sn = 93.8% | 91.6-97.8% |
|  | Negative | 24 | 153 | Sp = 45.4% | 39.3-49.7% |
| **Bacteriuria**<br>**(>125/μL)\*\* [8]** | Positive | 349 | 205 | Sn = 90.9% | 88.1-93.8% |
|  | Negative | 35 | 132 | Sp = 39.2% | 32.2-42.4% |

\*\*according to normal reference value, Sn: sensitivity, Sp: specificity

This Receiver Operating Characteristic (ROC) curve analyzes the diagnostic performance of UF BACT-count and WBC-count in distinguishing between conditions, likely indicating the presence or absence of a bacterial UTI. The UF BACT-count (green curve) demonstrates superior performance with an Area Under the Curve (AUC) of 0.85 and an optimal cut-off value of 975.4/µL. In contrast, the UF-WBC-count (blue curve) shows a lower diagnostic accuracy, indicated by an AUC of 0.69 and a cut-off value of 82.05/µL (Fig 3). A further ROC analysis based on sex indicated consistent diagnostic accuracy regardless of sex (S1 Fig). The diagnostic performance of the UF method in detecting bacteria and leukocytes at the optimal cutoff threshold in different sexes was presented in S1 Table.

**Figure 3.**
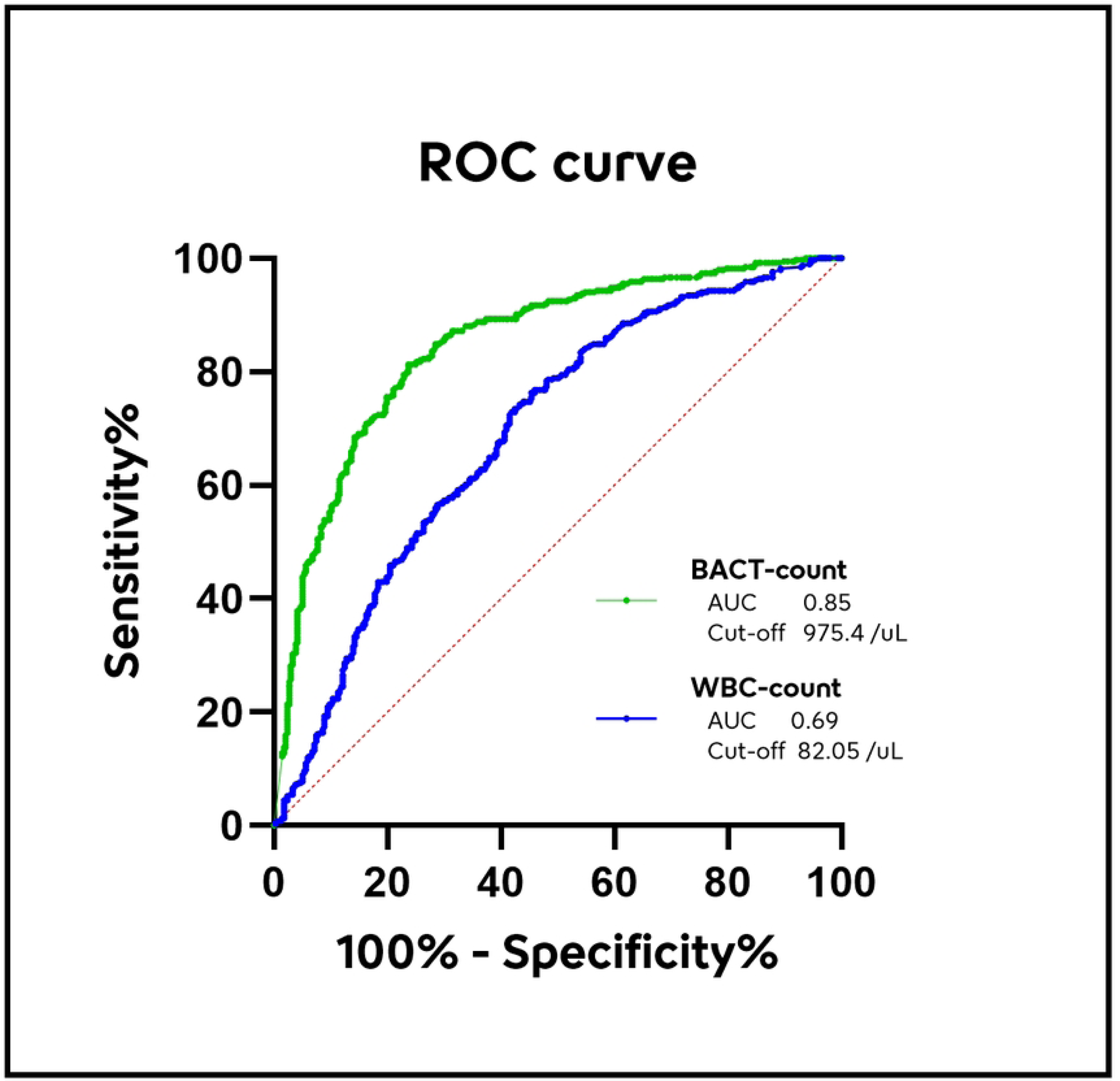
ROC analysis comparing BACT-count and WBC-count UF parameters in predicting UTI.

The UF UTI-Info. flag showed that out of 721 patients, 499 tested UTI positive and while 222 were not flagged as UTI positive (Table 7). Among those with a UTI flag, 343 had a positive urine culture (bacterial UTI), while 156 had a negative culture (non-bacterial UTI). Conversely, among those without a UTI flag, 41 had a positive culture and 181 had a negative culture. The UTI-Info. flag’s ability to detect UTI had an accuracy of 73%. These results used cutoffs of WBC >82.05/µL or bacteria >975.4/µL from this study.

**Table 7.** UF UTI-Info. Flag Validity Compared to Urine Culture (n=721) (Set up on WBC >82.05/µL or Bacteria >975.4/µL)

| UF UTI-Info. Flag | Urine Culture Positive | Urine Culture Negative | Total |
| --- | --- | --- | --- |
| Positive | 343 | 156 | 499 |
| Negative | 41 | 181 | 222 |
| **Total** | **384** | **337** | **721** |

### Evaluation of the BACT-Info. Flag in Relation to Urine Culture and Presumptive Gram Staining in UTIs

Among 312 samples analyzed, 40 cases were correctly identified as Gram-positive by the UF system, while 73 were misclassified as Gram-positive despite being sterile (Table 8). The observed agreement between UF flag and culture for Gram-positive bacteria was 66.67%, with a Kappa value of 0.216 (95% CI: 0.107–0.324), indicating only fair agreement and limited accuracy in identifying Gram-positive infections.

**Table 8.**
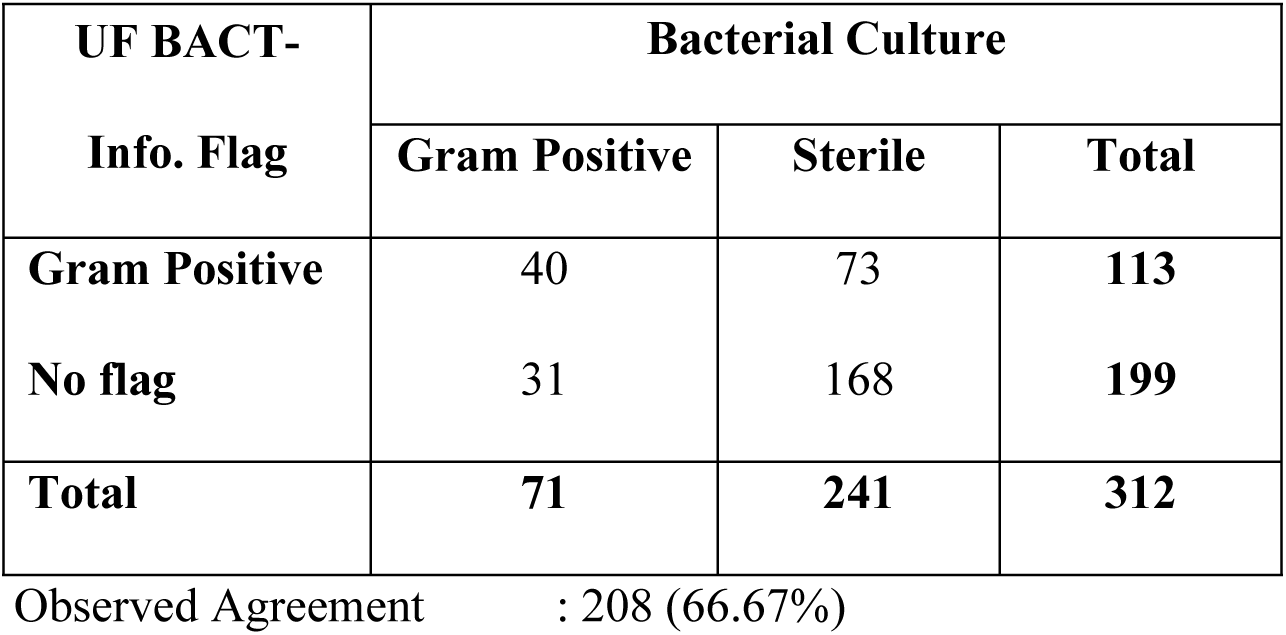
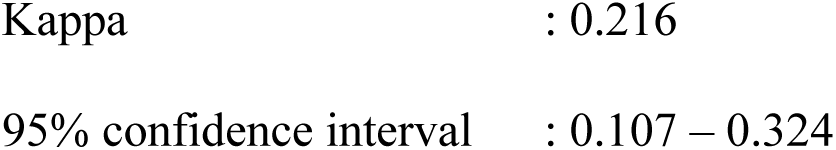
Comparison of Gram-Positive Results Between BACT-Info. and Bacterial Culture.

In contrast, table 9 presents a comparison for Gram-negative bacteria. Out of 315 samples, 104 were correctly flagged as Gram-negative, and only 12 sterile samples were misclassified as such. The observed agreement reached 86.35%, with a Kappa value of 0.716 (95% CI: 0.638–0.794), reflecting substantial agreement. This suggests that the UF-5000/4000 method is more reliable in detecting Gram-negative bacteria compared to Gram-positive ones in urinary tract infection screening.

**Table 9.** Comparison of Gram-Negative Results Between BACT-Info. and Bacterial Culture.

| UF BACT-Info. Flag | Bacterial Culture |  |  |
| --- | --- | --- | --- |
|  | Gram Negative | Sterile | Total |
| Gram Negative | 104 | 12 | **116** |
| No flag | 31 | 168 | **199** |
| **Total** | **135** | **180** | **315** |

Observed Agreement : 272 (86.35%)
Kappa : 0.716
95% confidence interval : 0.638 – 0.794

The comparison between BACT-Info. and presumptive Gram staining shows that the UF-5000/4000 demonstrates stronger performance in detecting Gram-negative bacteria than Gram-positive ones. For Gram-positive detection (Table 10), the observed agreement was 74.1%, with a moderate kappa value of 0.401 (95% CI: 0.303–0.514). In contrast, for Gram-negative detection (Table 11), the observed agreement was higher at 87.5% with a high kappa value of 0.721 (95% CI: 0.691–0.853), indicating substantial agreement. These results suggest that the UF-5000/4000 is consistently more reliable for identifying Gram-negative bacteria based on BACT-Info. flag compared to Gram-positive identification. The overall comparison results between BACT-Info. and either the Gram type determined from urine culture or the presumptive microscopic Gram type of urine are shown in S3 and S4 Tables.

**Table 10.** Comparison of Gram-Positive Results Between BACT-Info. and Presumptive Microscopic Gram Type.

| UF BACT-Info.<br>Flag | Presumptive Gram Type |  |  |
| --- | --- | --- | --- |
|  | Gram Positive | Sterile | Total |
| Gram Positive | 53 | 57 | 110 |
| No flag | 19 | 165 | 184 |
| Total | 72 | 222 | 294 |

Observed Agreement : 218 (74.1%)
Kappa : 0.401
95% confidence interval : 0.303 – 0.514

**Table 11.** Comparison of Gram-Negative Results Between BACT-Info. and Presumptive Microscopic Gram Type.

| UF BACT-Info.<br>Flag | Presumptive Gram Type |  |  |
| --- | --- | --- | --- |
|  | Gram Negative | Sterile | Total |
| Gram Negative | 86 | 17 | 103 |
| No flag | 19 | 165 | 184 |
| Total | 105 | 182 | 287 |

Observed Agreement : 251 (87.5%)
Kappa : 0.721
95% confidence interval : 0.691 – 0.853

## Discussion

Urine flow cytometry (UF) demonstrates significant potential for improving the efficiency and effectiveness of UTI diagnosis and management. Our findings, consistent with prior research [7, 9–11], indicate that UF-5000/4000 BACT-Info. flag reliably predicts bacterial Gram type, particularly for Gram-negative organisms. Specifically, we observed good agreement (Kappa = 0.716) between UF Gram-negative flags and culture results, notably reducing the use of broad-spectrum empirical antibiotics. There was also a good agreement between the UF no-flag category and sterile culture results, potentially reducing the need for unnecessary urine cultures by an estimated 59.1% (199 out of 337 sterile culture results). This aligns with Ren et al. report of a 61% reduction in unnecessary cultures and Pieretti et al. observation of up to 24-hour faster antibiotic initiation for negative urine results [7, 10]. While Gram-positive prediction showed lower agreement (Kappa = 0.216), consistent with moderate accuracy (sensitivity 0.70-0.80) reported by Wang et al., the strong positive predictive value (PPV = 0.93) of the Gram-negative flag for bacterial counts exceeding 1367/µL (as per Wang et al.) allows for rational empiric Gram-negative antibiotic initiation [12].

The ability of UF scattergrams to differentiate Gram-positive and Gram-negative bacteria up to 24 hours earlier than culture results, as highlighted by Enko et al. and supported by Yasutake et al. regarding scattergram patterns, offers a critical advantage for guiding early targeted therapy [11, 13]. This is further substantiated by Ozawa et al. and Yang et al., who noted that bacterial count and dominant scattergram angle predict Gram type, facilitating antibiotic initiation and selection before urine culture results are available [14, 15]. The consistency of UF results with microscopic leukocyturia also supports its utility for initiating empiric treatment.

Importantly, diagnostic accuracy may be further improved by incorporating manual verification techniques such as the 30-degree line rule, which evaluates scattergram angle patterns to distinguish true bacteriuria from debris or non-bacterial interference. Christy et al. demonstrated that combining automated bacterial flagging with targeted manual scattergram assessment enhances diagnostic performance, suggesting that laboratories adopting a hybrid automated–manual approach could achieve further gains in accuracy without substantial increases in workload [9]. Future studies integrating the 30-degree rule into UF screening algorithms may therefore help optimize interpretation and strengthen clinical decision-making. However, such a hybrid approach may be impractical for high throughput clinical laboratories with large sample volumes, and this may therefore be best reserved for selected cases where additional clarification is needed.

Despite its benefits, the diagnostic performance of UF for UTI predictions vary across studies due to differing cut-off values, diagnostic criteria, prevalence, and population characteristics. This underscores the need for each laboratory to establish and regularly validate optimal local cut-offs. Our evaluation of UF-5000/4000 revealed that WBC count did not significantly improve diagnostic accuracy, unlike bacterial count, which showed superior performance in UTI screening, consistent with Wang et al. study [12]. While WBC count and nitrite had minimal impact on screening validity in Wang’s study, the development of nomograms and decision trees based on routine urine parameters shows promise for enhancing screening validity.

In our cohort, when using a WBC cut-off of >82.05/μL or a bacterial count threshold >975.4/μL, the UF UTI-Info. flag achieved high sensitivity (89%) and NPV (82%), making it a reliable rule-out tool for infection. Although specificity was moderate (54%), the low negative likelihood ratio (0.20) indicates a substantial reduction in the positive post-test probability of UTI in UF-negative cases (false negative). This performance profile mirrors the characteristics desirable in a high-throughput screening tool, allowing clinicians to safely defer culture in low-risk patients while promptly initiating empiric therapy in high-probability cases. These findings support the strategic implementation of UF screening to reduce culture burden, avoid overtreatment, and guide antimicrobial stewardship efforts, particularly in high-volume or resource-constrained settings.

Based on these results and supporting evidence from the literature, we propose the following guideline algorithm for the screening and management of suspected UTIs, particularly to anticipate false negative UTI-Info. flag, integrating urinalysis and urine flow cytometry parameters (Fig 4). This guideline aims to assist clinicians in making timely and appropriate UTI treatment decisions.

**Figure 4.**
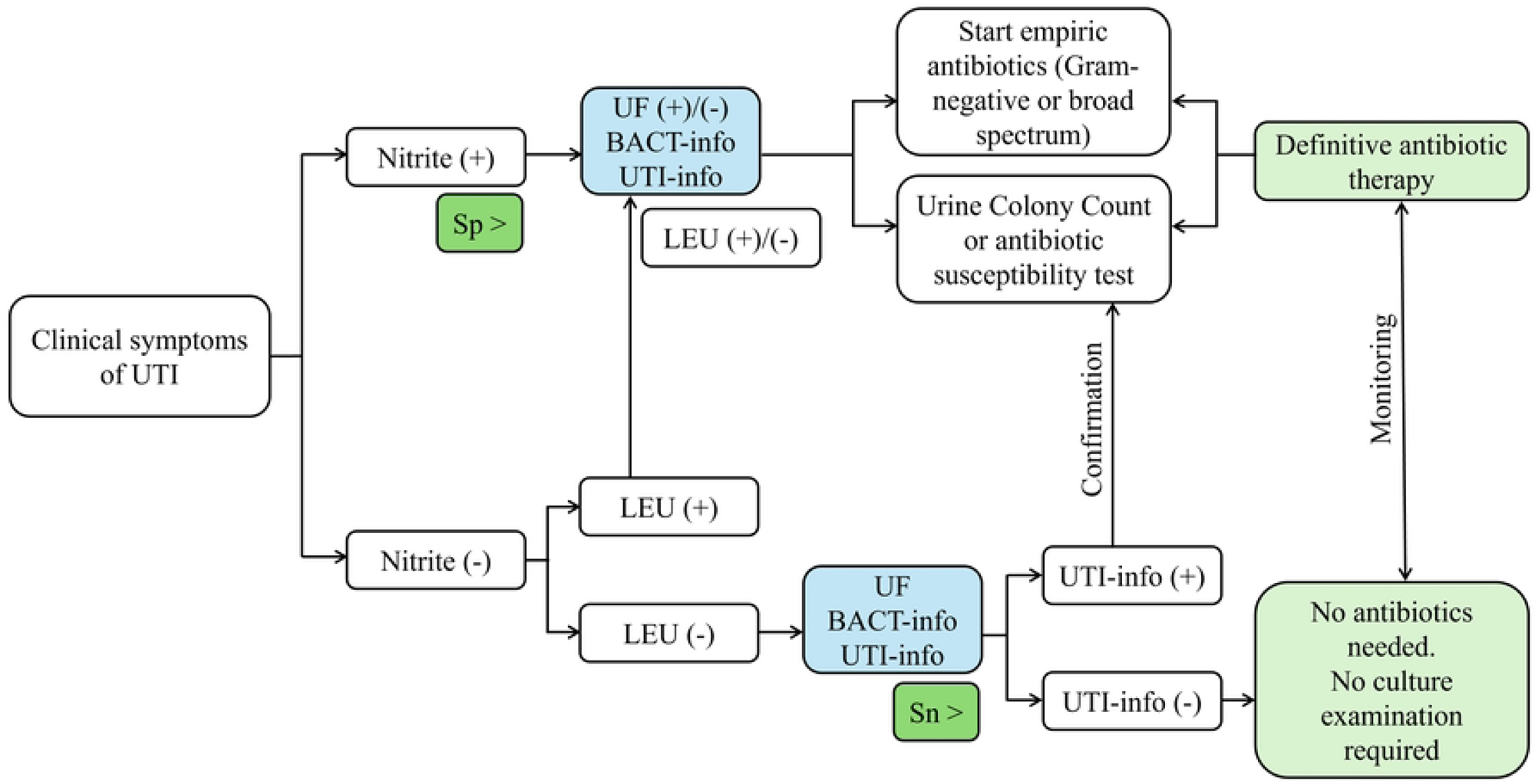
Proposed algorithm for screening and management of clinically suspected urinary tract infections (UTIs), integrating nitrite, leukocyte esterase, and urine flow cytometry (UF) results. The flowchart outlines decision pathways for initiating empiric antibiotics, performing urine culture, and ruling out UTI based on combined test findings.

The proposed algorithm depicted on Fig 4 primarily uses nitrite test results as the initial screening step for clinically suspected UTIs. As an initial screening step, nitrite test results are used due to their high specificity. If the nitrite result is positive, empiric antibiotic therapy can be initiated immediately, regardless of leukocyturia or UF results. Antibiotic selection can then be guided by UF “BACT-Info.” flag: a Gram-negative flag supports the use of narrow-spectrum antibiotics, while a Gram-positive flag may warrant broad-spectrum coverage. In outpatient settings with uncomplicated UTIs, urine culture may not be necessary, and treatment efficacy can be monitored through clinical symptoms and follow-up urinalysis. Conversely, for inpatients or cases involving complications, recurrence, or comorbidities, urine culture (urine colony count and antibiotic susceptibility test) remains essential to confirm pathogen identity and optimize antibiotic choice.

If the nitrite test is negative, a UTI cannot be ruled out based on this result alone due to its limited sensitivity. In such cases, leukocyturia and UF findings become critical. A positive leukocyturia result should prompt the same treatment considerations as a positive nitrite test. If leukocyturia is also negative, UF flag can assist further: a UTI-Info. flag still warrants proceeding with treatment, while a negative UTI-Info. flag, when combined with negative nitrite and leukocyturia results (a “triple-negative” profile), strongly suggests the absence of a UTI. In these cases, neither antibiotics nor urine culture are indicated. A limitation of this study is the non-inclusion of non-bacterial UTIs; therefore, caution is advised when evaluating cases caused by other urinary pathogens.

## Conclusion

Overall, the flow cytometry method providing UTI-Info. and BACT-Info. flags combined with urinalysis parameters (nitrite and leukocytes), presents a robust framework for developing UTI screening and diagnostic algorithms. This algorithm offers a valuable pathway for rapid prediction, particularly of common Gram-negative pathogens. Prompt initiation of empiric antibiotics is crucial for preventing complications such as bacteremia or urosepsis and improving clinical outcomes, and the strong agreement of Gram-negative BACT-Info. flag with presumptive Gram staining further supports reducing the reliance on microscopic examination. For cases where UF does not indicate UTI but clinical suspicion remains high (e.g., positive nitrite or leukocytes), conventional urine culture remains a critical safeguard.

## Data Availability

All relevant data are within the manuscript and its Supporting Information files.

## Supporting information

**S1 Figure. ROC Analysis Comparing BACT-count and WBC-count UF Parameters in Predicting UTI across general, female, and male populations**. When patients were further categorized based on sex, for both BACT-count (left) and WBC-count (right), the curves for general, female and male populations largely overlap, indicating consistent diagnostic accuracy regardless of sex. However, the BACT-count consistently demonstrates superior performance compared to the WBC-count, with its curve positioned further from the line of no discrimination, suggesting it is a more accurate diagnostic marker.

**S1 Table 1. Microorganisms Identified from Positive Culture Results (n=425).** The most commonly isolated pathogen in this study was *Escherichia coli*, accounting for 37.41% of all isolates, followed by *Klebsiella pneumoniae* (11.76%) and *Burkholderia cepacia* (6.12%). Other notable bacteria included *Enterococcus faecalis* and *Pseudomonas aeruginosa* (each 5.88%), while fungal isolates such as *Candida albicans* (2.82%) and *Candida tropicalis* (1.88%) were also detected. Less frequent pathogens included *Acinetobacter baumannii* (2.35%), *Staphylococcus epidermidis* (2.12%), and *Proteus mirabilis* (1.88%). Additionally, 6.35% of isolates were classified as other Gram-positive bacteria, 11.53% as other Gram-negative bacteria, and 4.00% as other fungi.

**S2 Table. Diagnostic Performance of the UF Method in Detecting Bacteria and Leukocytes at the Optimal Cutoff Threshold**. For the general population, a BACT-count cutoff of 975.4/µL yielded 81.3% sensitivity and 76.3% specificity. Among males, the optimal BACT-count cutoff was 1008/µL (80.0% sensitivity, 80.1% specificity), while for females it was 967.7/µL (81.9% sensitivity, 72.9% specificity). For WBC-count, the general population cutoff was 82.05/µL with 72.7% sensitivity and 58.5% specificity. In males, a lower cutoff of 51.4/µL achieved higher sensitivity (81.6%) but moderate specificity (53.2%), whereas in females, a cutoff of 83.0/µL yielded 71.8% sensitivity and 59.7% specificity. These findings suggest good diagnostic potential of UF parameters, particularly BACT-count, across different subgroups.

**S3 Table. Distribution of BACT-Info. UF-5000/4000 Flag Results Based on Gram Type of Urine Culture Results**. Among the 721 total samples, urine culture identified 318 as Gram-negative, 59 as Gram-positive, 7 as mixed bacterial, and 337 as sterile. The UF system flagged 118 samples as Gram-negative, of which 104 matched the culture results. It flagged 172 as Gram-positive, with only 40 correctly identified. The mixed flag was applied to 143 cases, though only 5 were confirmed as mixed. A total of 199 samples showed no flag, most of which (168) were truly sterile in urine culture. This distribution suggests that while the UF system has a relatively good ability to flag Gram-negative bacteria, its accuracy is lower for detecting Gram-positive and mixed-Gram bacterial UTIs.

**S4 Table. Distribution of BACT-Info. UF-5000/4000 Flag Results Based on Presumptive Microscopic Gram Type of Urine**. When the UF-5000/4000 system results were compared to the presumptive microscopic Gram typing, 86 out of 240 culture-confirmed Gram-negative cases were correctly flagged by the UF as Gram-negative, while the remaining cases were misclassified under different categories (table 11). Similarly, out of 117 presumptive microscopic Gram-positive cases, 53 were correctly identified by the UF system, and 64 were misclassified as various categories. Mixed and non-classifiable BACT-Info. flag results also showed varied distribution with the UF system identifying 24 mixed infections, consistent with the presumptive Gram results, and 70 non-classified results categorized as having no bacteria found. The “No flag” category in the UF system had the highest number of sterile cases (165 out of 199), indicating the system’s tendency not to flag when bacterial presence was low or absent. This category also showed the highest agreement with the absence of bacteria in presumptive Gram results.

## References

1. Yashir M, Apriani A. Variasi bakteri pada penderita infeksi saluran kemih (ISK). Jurnal Media Kesehatan. 2019; 12(2): 102–109. 10.33088/jmk.v12i2.441

2. Medina M, Castillo-Pino E. An introduction to the epidemiology and burden of urinary tract infections. Ther Adv Urol. 2019;11:1756287219832172. doi:10.1177/1756287219832172. PMID: 31105774; PMCID: PMC6502976.

3. Salam MA, Al-Amin MY, Salam MT, Pawar JS, Akhter N, Rabaan AA, Alqumber MAA. Antimicrobial Resistance: A Growing Serious Threat for Global Public Health. Healthcare (Basel). 2023;11(13):1946. doi: 10.3390/healthcare11131946. PMID: 37444780; PMCID: PMC10340576.

4. Claeys KC, Blanco N, Morgan DJ, Leekha S, Sullivan KV. Advances and Challenges in the Diagnosis and Treatment of Urinary Tract Infections: the Need for Diagnostic Stewardship. Curr Infect Dis Rep. 2019;21(4):11. doi: 10.1007/s11908-019-0668-7. PMID: 30834993.

5. Schmiemann G, Kniehl E, Gebhardt K, Matejczyk MM, Hummers-Pradier E. The diagnosis of urinary tract infection: a systematic review. Dtsch Arztebl Int. 2010;107(21):361–7. doi: 10.3238/arztebl.2010.0361. Epub 2010 May 28. PMID: 20539810; PMCID: PMC2883276.

6. Broeren MA, Bahçeci S, Vader HL, Arents NL. Screening for urinary tract infection with the Sysmex UF-1000i urine flow cytometer. J Clin Microbiol. 2011;49(3):1025–9. doi: 10.1128/JCM.01669-10. Epub 2011 Jan 19. PMID: 21248088; PMCID: PMC3067737.

7. Ren C, Wu J, Jin M, Wang X, Cao H. Rapidly discriminating culture-negative urine specimens from patients with suspected urinary tract infections by UF-5000. Bioanalysis. 2018;10(22):1833–1840. doi: 10.4155/bio-2018-0175. Epub 2018 Oct 8. PMID: 30295053.

8. Manoni F, Fornasiero L, Ercolin M, Tinello A, Ferrian M, Hoffer P, Valverde S, Gessoni G. Cutoff values for bacteria and leukocytes for urine flow cytometer Sysmex UF-1000i in urinary tract infections. Diagn Microbiol Infect Dis. 2009;65(2):103–7. doi: 10.1016/j.diagmicrobio.2009.06.003. PMID: 19748419.

9. Christy P, Sidjabat HE, Lumban Toruan AA, Moses EJ, Mohd Yussof N, Puspitasari Y, Fuadi MR, Aryati, Marpaung FR. Comparison of Laboratory Diagnosis of Urinary Tract Infections Based on Leukocyte and Bacterial Parameters Using Standardized Microscopic and Flow Cytometry Methods. Int J Nephrol. 2022;2022:9555121. doi: 10.1155/2022/9555121. PMID: 35669495; PMCID: PMC9167024.

10. Pieretti B, Brunati P, Pini B, Colzani C, Congedo P, Rocchi M, Terramocci R. Diagnosis of bacteriuria and leukocyturia by automated flow cytometry compared with urine culture. J Clin Microbiol. 2010;48(11):3990–6. doi: 10.1128/JCM.00975-10. Epub 2010 Aug 25. PMID: 20739491; PMCID: PMC3020858.

11. Enko D, Stelzer I, Böckl M, Schnedl WJ, Meinitzer A, Herrmann M, Tötsch M, Gehrer M. Comparison of the reliability of Gram-negative and Gram-positive flags of the Sysmex UF-5000 with manual Gram stain and urine culture results. Clin Chem Lab Med. 2020;59(3):619–624. doi: 10.1515/cclm-2020-1263. PMID: 33068381.

12. Wang H, Han FF, Wen JX, Yan Z, Han YQ, Hu ZD, Zheng WQ. Accuracy of the Sysmex UF-5000 analyzer for urinary tract infection screening and pathogen classification. PLoS One. 2023;18(2):e0281118. doi: 10.1371/journal.pone.0281118. PMID: 36724192; PMCID: PMC9891513.

13. Yasutake Y, Higuchi M, Oda S, Tamura Y, Shimadu T, Kobayashi H, Hanaoka E. Comparisons of the Bact scattergram pattern by fully automated integrated urine analyzer UX-2000 and microscopic examination results using gram stain. Sysmex Journal International. 2013; 23(1). Available from: https://www.sysmex.co.jp/en/products_solutions/library/journal/vol23_no1/vol23_1_08.pdf

14. Ozawa H, Yajima N, Kobayashi H. Estimation of the causative bacterial group from bacterial scattergrams of the fully automated urine particle analyzer UF-1000i. Sysmex Journal International. 2012; 22(1). Available from: https://www.sysmex.co.jp/en/products_solutions/library/journal/vol22_no1/vol22_1_06.pdf

15. Yang CC, Yang SS, Hung HC, Chiang IN, Peng CH, Chang SJ. Rapid differentiation of cocci/mixed bacteria from rods in voided urine culture of women with uncomplicated urinary tract infections. J Clin Lab Anal. 2017;31(5):e22071. doi: 10.1002/jcla.22071. Epub 2016 Nov 15. PMID: 27859671; PMCID: PMC6817067.

